# Cerebrospinal fluid transcriptional immune pathways linked to survival in HIV-associated tuberculous meningitis

**DOI:** 10.1101/2025.11.05.25339630

**Authors:** Martineau Louine, Ravi Dandekar, Sumanth P. Reddy, Mary C. Karalius, Greer Waldrop, Shinyin Wang, Jane Gakuru, Sarah Kimuda, Timothy Mugabi, Abdu K Musubire, Enock Kagimu, Mahsa Abassi, Mable Kabahubya, Darlisha A. Williams, Hoang Van Phan, Biyue Dai, Maham Zia, Kelsey C. Zorn, Camille Fouassier, Chloe Gerungan, Pedro S. Marra, Caleb P Skipper, Nathan C. Bahr, Charles R. Langelier, Fiona V. Creswell, David R. Boulware, David B. Meya, Michael R. Wilson

## Abstract

**BACKGROUND:** TB meningitis (TBM) has up to 50% mortality in people living with HIV. We investigated differences in cerebrospinal fluid (CSF) host immune responses associated with acute mortality.

**METHODS:** We enrolled a prospective cohort of adults with definite, probable and possible HIV-related TBM, without evidence of co-infection, in Kampala, Uganda. We performed metagenomic next-generation sequencing (mNGS) of bulk CSF RNA to profile host gene expression and exclude participants with co-infecting or alternate central nervous system pathogens. We then assessed host differential gene expression based on 14-day mortality within the refined cohort.

**RESULTS:** Among the 110 patients included in the transcriptomic analysis, 22.7% (n=25) died within 14 days of enrollment. More than 2000 genes were differentially expressed in the CSF based on 14-day mortality (adjusted p-value < 0.05). Genes upregulated in TBM-survivors included genes related to T-cell receptor signaling (*LCK, FYN, LAT*), T-cell survival and differentiation (*IL7, CD27, IL12RB1*), B-cell receptor signaling (*BCR, CD81, PLCG2*), and TNF signaling. Survivors demonstrated downregulation in the neutrophil chemoattractant gene *CXCL1*, and classical complement pathway genes *C4A* and *C4B*.

**CONCLUSIONS:** A regulated immune response in which T-cell, B-cell, and microglial signaling are upregulated, but also in which certain neutrophil and complement genes are downregulated, was associated with short-term TBM survival in this population with HIV-related TBM. This finding provides context for the nuanced immunologic response and suggests that certain targeted immunomodulatory agents may be more effective as adjunctive therapy in HIV-related TBM compared to broad spectrum agents such as glucocorticoids.

## INTRODUCTION

Approximately one-quarter of the world’s population is infected with *Mycobacterium tuberculosis* (*Mtb*), and over 10 million people each year develop active tuberculosis (TB).^1^ TB remains the leading infectious cause of death worldwide, responsible for 1.25 million deaths in 2023, including 161,000 deaths among people living with HIV.^1^ Tuberculous meningitis (TBM) is a devastating manifestation of TB, with a mortality rate of approximately 50% among people living with HIV.^2^ In 2018, the National Institutes of Health convened a workshop to identify knowledge gaps impeding the management of TBM, and identified two main culprits: 1) the lack of gold-standard diagnostic testing that is both sensitive and specific for detecting *Mtb* in the cerebrospinal fluid (CSF), and 2) a poor understanding of TBM immunopathogenesis.^3^

To overcome diagnostic limitations, recent efforts have focused on improving nucleic-acid-based testing for *Mtb* in the CSF. Both targeted methods, such as TB polymerase chain reaction (PCR), and agnostic approaches, such as metagenomic next-generation sequencing (mNGS), can increase the likelihood of diagnosing TBM.^4–6^ However, questions remain regarding TBM pathogenesis, including the degree to which mortality is driven by *Mtb* itself versus the exuberant immune response to the infection.^7,8^ For example, although adjunctive dexamethasone confers a survival benefit in TBM, a sub-group analysis found that the trend towards a survival benefit was smaller and not statistically significant among patients living with HIV (Hazard Ratio (HR) = 0.86; 95%CI: 0.52-1.41).^9^ This prompted the Adjunctive Corticosteroids for Tuberculous Meningitis in HIV-positive adults (ACT HIV) trial, which similarly found that this trend towards mortality benefit for adjunctive dexamethasone was nonsignificant (HR = 0.85; 95%CI: 0.66 to 1.10).^10^ These trials illustrate that the differences in the survival benefit of adjunctive immunosuppressive therapy in various patient populations is at least in part due to differing central nervous system (CNS) immune responses.^8,11^ Furthermore, although neither trial found a statistically significant mortality benefit with adjunctive glucocorticoid therapy, the *trend* towards a benefit suggests that a subset of TBM patients living with HIV may still benefit from adjunctive immunosuppressive therapy.

This study seeks to better understand the nuances of the CNS host response to *Mtb* among patients living with HIV, and whether differences are associated with short-term mortality. We analyzed a well-characterized clinical cohort of Ugandan adults living with HIV and diagnosed with possible, probable, or definite meningitis via routine clinical testing.^12–14^ CSF mNGS was first utilized to refine the cohort to those with probable or definite TBM and without co-infecting or alternate CNS pathogens. Differential gene expression analysis of bulk CSF transcriptomes was then performed to identify potential biological mechanisms associated with 14-day mortality among patients living with HIV and comorbid TBM.

## RESULTS

### Cohort refinement via CSF mNGS for pathogen detection

From 2018-2023, 163 adults who were hospitalized in Uganda with possible, probable, or definite TBM (as defined by the Uniform Case Definition)^12^ were enrolled (**Figure 1a**). After exclusion of participants with negative or unknown HIV testing (n=20) or with greater than 3 days of anti-TB treatment prior to lumbar puncture (n=10), the remaining 133 participants were included in our study and underwent CSF mNGS testing. The average sequencing depth was 73 million RNA reads per sample (IQR: 26 million – 148 million), and after computational filtering and removal of host reads, an average of 4,212,604 RNA reads passed filters (IQR: 642,428 – 4,990,796). CSF mNGS detected *Mtb* in 75.5% (40/53) of participants diagnosed with definite TBM via Xpert MTB/Rif Ultra or culture. CSF mNGS detected *Mtb* in 7 participants with probable TBM (n=63) and 2 participants with possible TBM (n=17), allowing for re-classification of these cases as definite TBM for the purposes of the transcriptomic analysis (**Figure 1b**). Among the participants with possible TBM who were excluded due to lack of *Mtb* detection, two clinically significant alternate neurologic infections (herpes simplex-1 virus and varicella zoster virus) were detected by mNGS. Three cases of definite TBM were excluded due to detection of *Toxoplasma gondii* co-infection by CSF mNGS. Five cases of probable TBM were excluded due to detection of *T. gondii* (n=5) and *Cryptococcus neoformans* (n=1, occurred in tandem with *T. gondii,* **Figure 1c**).

**Figure 1:**
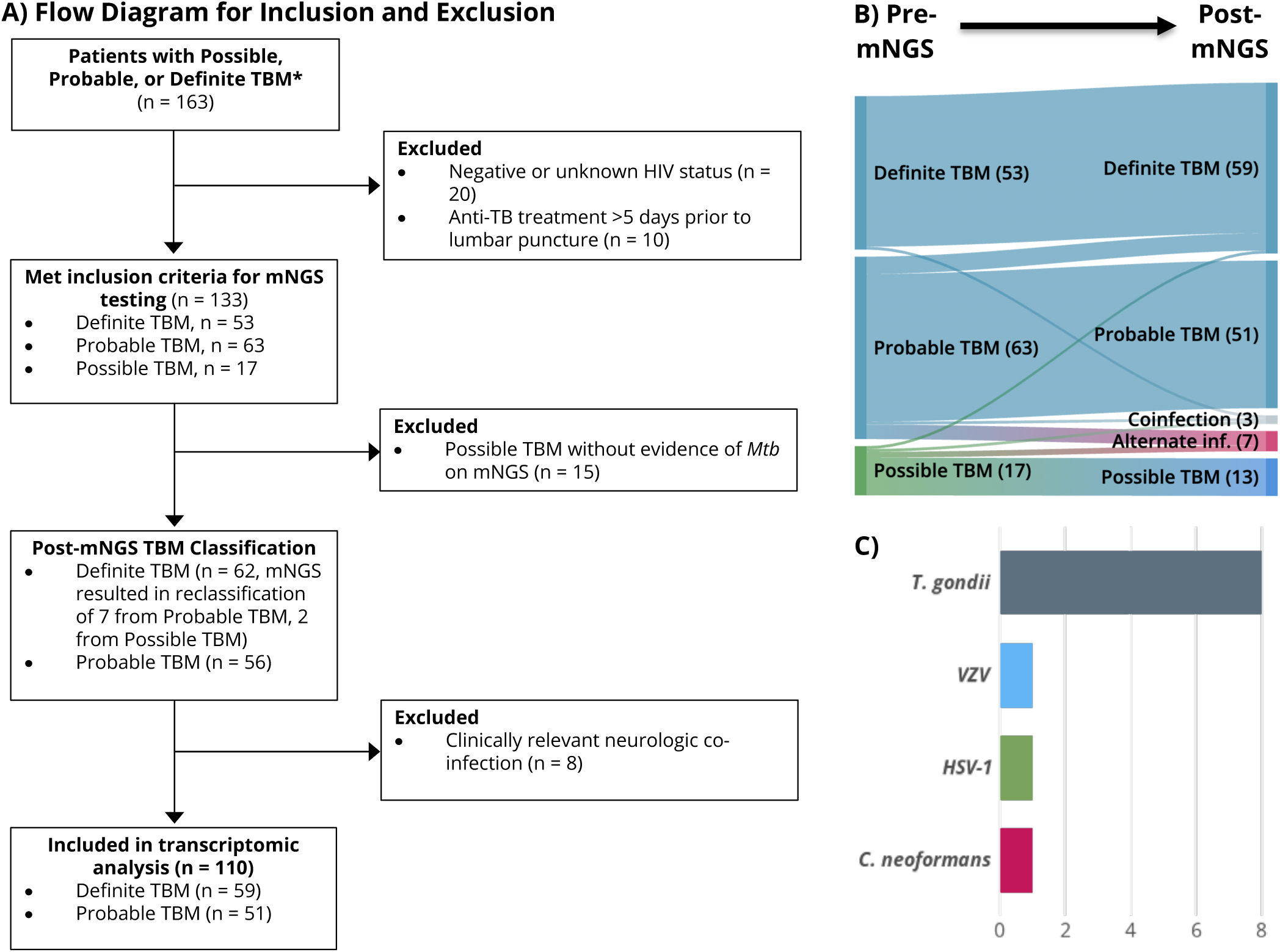
Study overview. **A)** Flow diagram describing the inclusion and exclusion process for CSF mNGS testing and the final transcriptomic analysis. **B)** Sankey diagram depicting how CSF mNGS impacted the diagnostic recharacterization for the 133 patients with possible, probable, and definite TBM who underwent CFS mNGS. **C)** CNS alternate infections and co-infections detected by CSF mNGS.

Following cohort refinement based on CSF mNGS results, 110 participants living with HIV and with probable TBM (n=51, defined by the Uniform Case Definition)^12^ or definite TBM (n=59, defined by microbiological confirmation on routine clinical testing or on CSF mNGS) without evidence of alternative or comorbid neurologic infections were included in the transcriptomic analysis.

### Cohort Demographics

Among the 110 participants, 48% (n=53) were female, the median age was 35 years (IQR: 30, 42), and the median CD4 count was 78 cells/μL (**Table 1**, IQR:38, 186). Thirty-eight percent (41/110) were receiving antiretroviral therapy (ART) at presentation with a median duration of therapy of 4 months (IQR:1, 6). Most participants presented with altered mental status (Glasgow Coma Scale score < 15, 83%, n=89/107). The median CSF white cell count was 45 cells/μL (n=105, IQR: <5, 200), and the median opening pressure was 170 mmH_2_O (n=89, IQR:120-240). Overall, 85 (78%) of the 110 participants were alive 14 days after study enrollment, and the 14-day survival was higher among males compared to females (86% vs 68%, p=0.024). There were no other statistically significant associations between baseline demographic and clinical characteristics and 14-day survival among this cohort (**Table 1**).

**Table 1.** Baseline characteristics by 14-day mortality status.

| Characteristic | n | Overall, n = 110<sup>1</sup> | n | Alive<sup>2</sup>, n = 85<sup>1</sup> | n | Dead, n = 25<sup>1</sup> | p-value |
| --- | --- | --- | --- | --- | --- | --- | --- |
| Age | 110 | 35 (30, 42) | 85 | 35 (30, 42) | 25 | 36 (28, 42) | 0.63 |
| Female sex | 110 | 53 (48%) | 85 | 36 (42%) | 25 | 17 (68%) | 0.024 |
| Diagnostic Category after mNGS |  |  |  |  |  |  | 0.79 |
| Definite TBM |  | 59 (54%) |  | 45 (53%) |  | 14 (56%) |  |
| Probable TBM |  | 51 (46%) |  | 40 (47%) |  | 11 (44%) |  |
| CD4 Count | 81 | 78 (38, 186) | 66 | 82 (46, 203) | 15 | 46 (24, 129) | 0.12 |
| ART<sup>3</sup> Groups | 108 |  | 84 |  | 24 |  | 0.22 |
| ART experienced but not currently receiving |  | 21 (19%) |  | 14 (17%) |  | 7 (29%) |  |
| ART naive |  | 46 (43%) |  | 35 (42%) |  | 11 (46%) |  |
| Currently on ART |  | 41 (38%) |  | 35 (42%) |  | 6 (25%) |  |
| Months on ART | 39 | 4 (1, 16) | 34 | 4 (1, 16) | 5 | 4 (3, 9) | 0.92 |
| GCS<sup>4</sup> score < 15 | 107 | 89 (83%) | 83 | 66 (80%) | 24 | 23 (96%) | 0.07 |
| Cerebrospinal Fluid Testing |  |  |  |  |  |  |  |
| White blood cells (cells/μL) | 105 | 45 (4, 200) | 80 | 55 (4, 205) | 25 | 4 (4, 160) | 0.43 |
| Protein (mg/dL) | 101 | 105 (60, 142) | 77 | 105 (73, 140) | 24 | 105 (59, 187) | 1.0 |
| Glucose (mg/dL) | 99 | 44 (22, 75) | 76 | 46 (23, 73) | 23 | 33 (20, 87) | 0.76 |
| Lactate (mmol/L) | 80 | 5.1 (3.2, 8.0) | 58 | 4.8 (2.8, 7.6) | 22 | 6.5 (3.3, 8.7) | 0.20 |
| Opening Pressure (mmH<sub>2</sub>O) | 89 | 170 (120, 240) | 68 | 178 (128, 233) | 21 | 150 (70, 240) | 0.490 |

<sup>1</sup>Median (IQR); n (%)<sup>2</sup>Includes patients who were discharged prior to hospital day 14.<sup>3</sup>Antiretroviral therapy<sup>4</sup>Glasgow Coma Scale

### Host transcriptomics and differential gene expression

The median non-zero protein-coding gene count in survivors was 8787 [IQR: 3058-14241] versus 3482 [IQR: 2351-8861] in non-survivors (p=0.02). Differential gene expression analysis revealed that 2190 genes were differentially expressed (Benjamini-Hochberg adjusted p-value < 0.05 and an absolute log-fold change ≥ 1.5) between 14-day TBM survivors and non-survivors, including 1770 upregulated genes and 420 downregulated genes (**Figure 2a)**. Common biological coefficient of variation (BCV) estimates ranged from 0.8 to 2.6 (**Supplementary Figure 1**).

**Figure 2:**
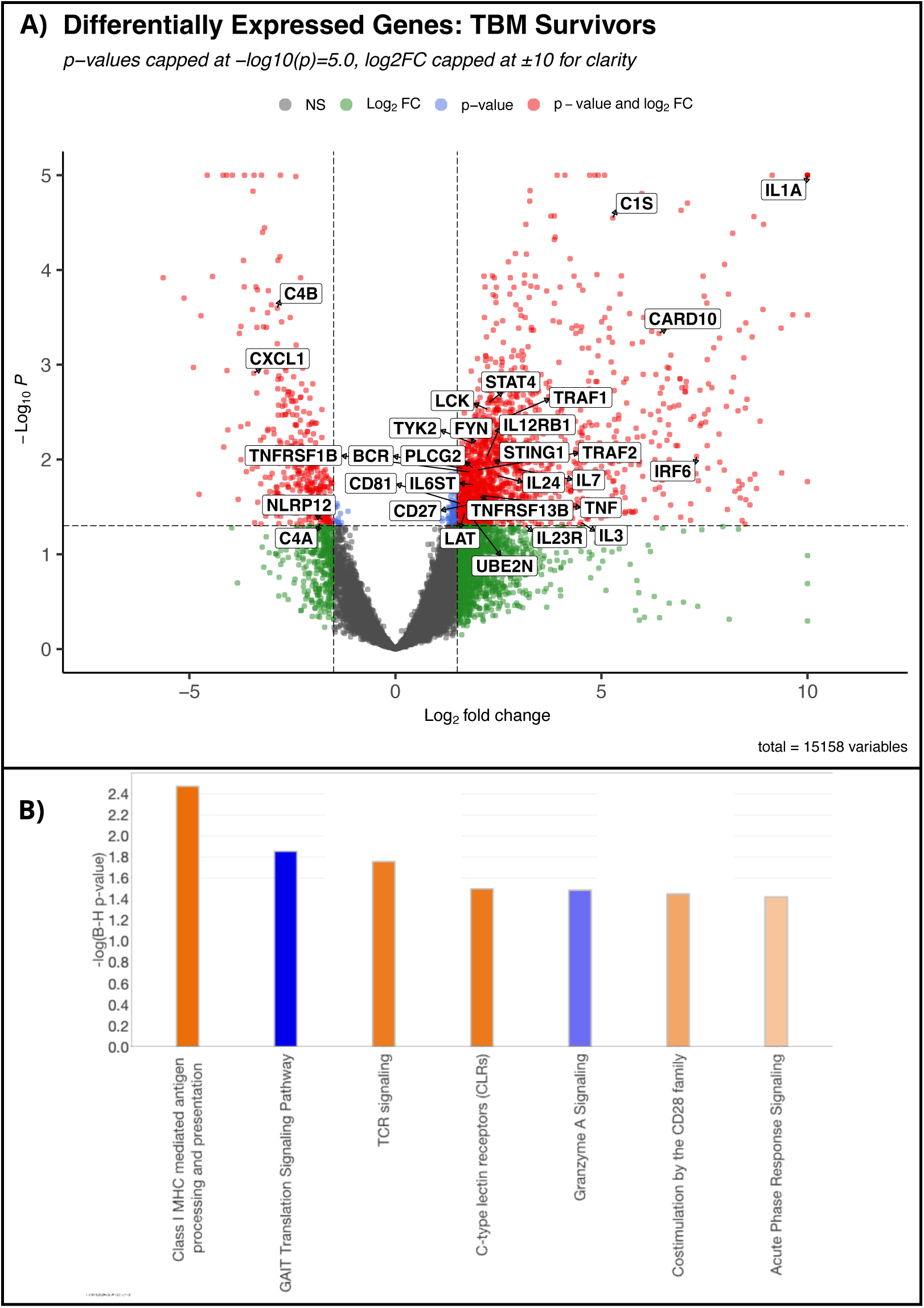
Differential expression of immune relevant genes and pathways based on 14-day mortality status in HIV-associated TBM. **A)** Volcano plot demonstrating upregulated and downregulated genes among 14-day survivors (NS: non-significant, FC: fold change). In comparison to patients who died before day 14, significantly upregulated proinflammatory genes included IL1A, TNF, and NF-kB pathway, and BCR signaling components, whereas IL-1RN and complement proteins C4A and C4B were downregulated among 14-day survivors. **B)** Ingenuity Pathway Analysis visualization of immunologically relevant pathways upregulated (orange) or downregulated (blue) among TBM survivors. MHC: Major histocompatibility complex; GAIT: interferon-γ-activated inhibitor of translation; TCR: T-cell receptor.

Transcriptomic analysis revealed that TBM survivors exhibited upregulation of multiple lymphoid immune signaling components. Expression of T-cell receptor (TCR) signaling genes was increased in survivors, including the proximal kinases LCK proto-oncogene (*LCK*) and FYN proto-oncogene (*FYN*), along with the adaptor protein linker for activation of T-cells (*LAT*). T-cell survival and differentiation genes also demonstrated increased expression among survivors, including interleukin 7 (*IL7*), *CD27*, interleukin-12 receptor subunit beta 1 (*IL12RB1*), and signal transducer and activator of transcription 4 (*STAT4*). B-cell signaling genes were similarly upregulated, including B-cell receptor (*BCR*), *CD81*, and phospholipase C gamma 2 (*PLCG2*).

Survivors also demonstrated upregulation of additional genes with functions in both innate and adaptative immune signaling including interleukin 1A (*IL1A*), tumor necrosis factor (*TNF*), and canonical NF-κB pathway components: caspase recruitment domain family member 10 (*CARD10*), ubiquitin conjugating enzyme E2 (*UBE2N*), and TNF receptor-associated factor 2 (*TRAF2*). Interferon signaling pathways were also enhanced, with increased expression of stimulator of interferon genes 1 (*STING1*), interferon regulatory factor 6 (*IRF6*), and tyrosine kinase 2 (*TYK2*). Conversely, survivors exhibited downregulation of interleukin 1 receptor antagonist (*IL1RN*), the neutrophil chemoattractant gene *CXCL1*, and classical complement pathway genes *C4A* and *C4B*.

**Figure 3:**
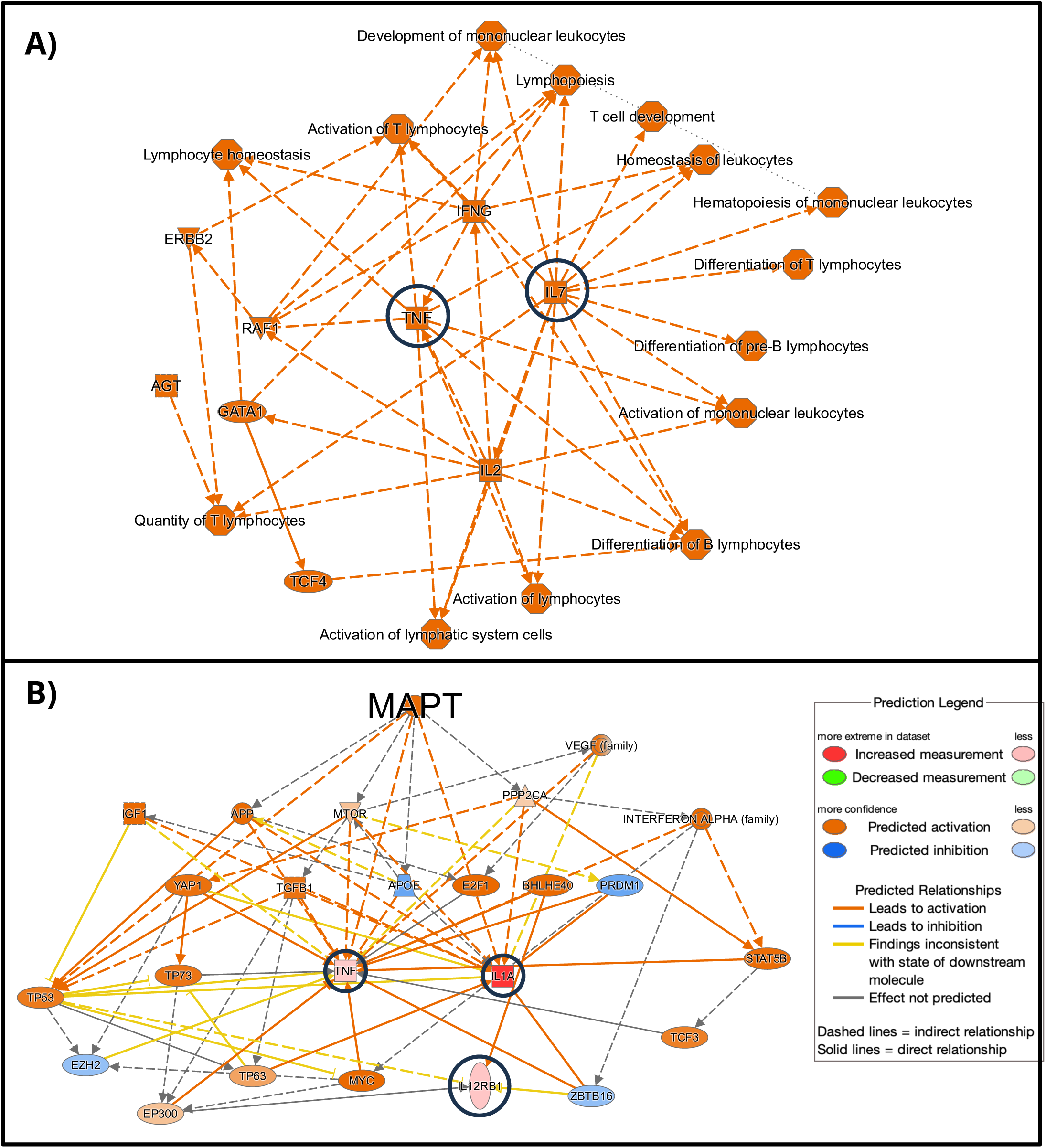
Mediators and Upstream Regulators of the anti-*Mtb* Immune Response. A) Graphical summary of the IPA analysis demonstrating the central role of TNF and IL7 in the function of B and T lymphocytes among survivors. Dashed lines indicate indirect interactions between the molecules and biological functions (hexagons). B) Network interaction plot of MAPT based on IPA upstream regulator analysis. MAPT indirectly activates several signaling molecules, leading to the downstream upregulation of several immunologic genes associated with survival, including TNF, IL1A, and IL12RB1 (black circles). Legend: square = cytokine, inverted triangle = kinase, horizontal oval = transcriptional regulator, vertical oval = transmembrane receptor, trapezoid = transporter, circle = other.

Unsupervised hierarchical clustering identified a subgroup (n=26) with elevated mortality (38% vs. 28% and 9%, p=0.02), lower CSF lactate (p=0.04), and a lower number of protein coding genes with nonzero counts (median 2,392 vs. 20,574 and 6,393, p < 0.001). There were no other significant differences between baseline characteristics between the clusters (**Supplemental Table 1**). The “hypoinflammatory” cluster with elevated mortality was characterized by global downregulation of both innate and adaptive immune pathways (**Figure 4a**), including antigen presentation genes, T-cell activation and trafficking genes, Th1 signaling genes, myeloid/macrophage activation markers, and interferon alpha and beta receptor subunit 1 (*INFNAR1,* **Figure 4b**). The hypoinflammatory cluster also demonstrated a mixed pattern of complement pathway gene expression, with downregulation of *C1QA* and *C1QC*, and upregulation of *C7*. Neutrophil-associated genes including *CXCL2* and Fc fragment of IgG receptor IIIb (*FCGR3B*) were also significantly downregulated. Within the hypoinflammatory cluster, survivors (n=16) demonstrated upregulation of TNF receptor superfamily members 1A and 1B (*TNFRSF1A*, *TNFRSF1B*), interferon gamma receptor 2 (*IFNGR2*), integrin subunit alpha L (*ITGAL*), interleukin 1 receptor accessory protein (*IL1RAP*), Fc fragment of IgE receptor Ig (*FCER1G*), and *CCL5,* in comparison to nonsurvivors (**Figure 4c**).

**Figure 4:**
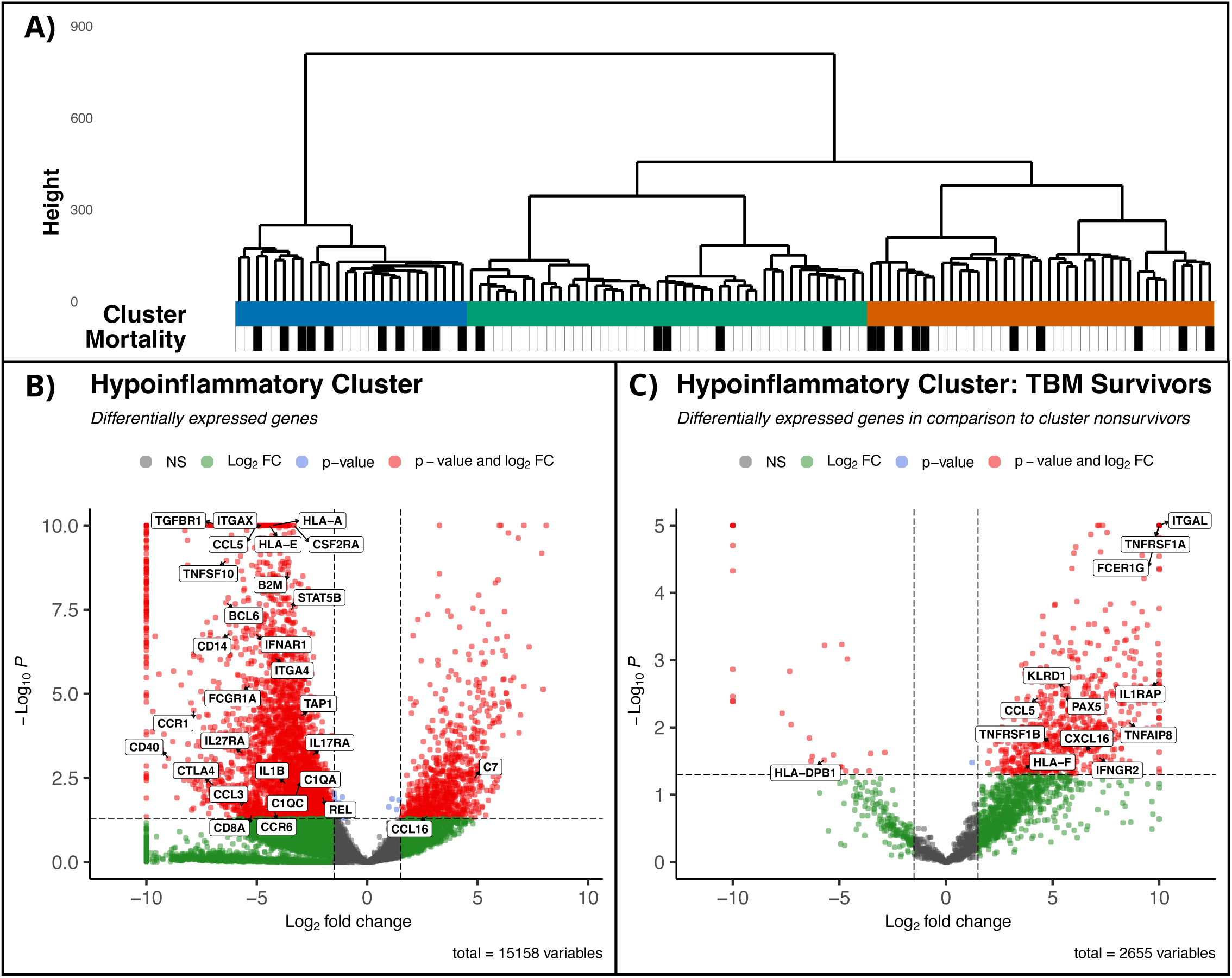
Unsupervised heirarchical clustering reveals a hypoinflammatory phenotype associated with increased mortality. **A)** Hierarchical clustering dendrogram of patients based on the top 1,000 most variable protein-coding genes. Height represents the Euclidean distance at which clusters merge. Three clusters were identified (blue = cluster 1, teal = cluster 2, orange = cluster 3), with black bars indicating non-survivors. Cluster 1 (blue) represents a hypoinflammatory phenotype with significantly elevated mortality (38% vs. 9% and 28%, p=0.02) and lower CSF lactate levels (p=0.04), and is characterized by **B)** global downregulation of genes involved in innate and adaptive immune pathways, compared to the other two clusters. **C)** Within the hypoinflammatory cluster, survivors demonstrated upregulation of genes related to TNF, interferon gamma, and IL1 signaling, among others.

### Gene pathway analysis

Ingenuity Pathway Analysis (Qiagen)^15^ was used to contextualize differentially expressed genes within known biological pathways and networks (**Figure 2b**). Inflammatory pathways significantly upregulated in TBM survivors included class I MHC-mediated antigen processing and presentation, TCR signaling, C-type lectin receptors (CLRs), CD28 family co-stimulation, and acute phase response signaling. In contrast, TBM survivors demonstrated significant downregulation of the interferon-γ-activated inhibitor of translation (GAIT translation) and granzyme A signaling pathways.

Upstream regulator analysis identified that microtubule associated protein tau (MAPT) was predicted to be activated among TBM survivors based on downstream target activation of several immunologic genes, including *TNF*, *IL1A*, and *IL12RB1*, and inhibition of *IL1RN* (**Figure 4b**). Other significant upstream immunologic regulators that were predicted to be activated based on downstream target activation included *TNF*, *IL7*, X-box binding protein 1 (*XBP1*), and transcription factor EB (*TFEB*). The graphical representation in **Figure 4a** highlights the multifaceted role of the upstream regulators *IL7* and *TNF* in the development, activation, differentiation, and homeostasis of T-cells, B-cells, and mononuclear cells.

## DISCUSSION

In this study, we define CSF transcriptional signatures associated with TBM survival in patients living with HIV. After rigorous cohort refinement using CSF mNGS to detect pathogens and minimize diagnostic misclassification, differential gene expression and pathway analysis demonstrated that short-term survival was associated with broad immune upregulation, but with some restraint of innate immune responses including complement genes and neutrophil recruitment. The hierarchical clustering analysis identified a subset of patients with a global hypoinflammatory phenotype and increased mortality, adding additional context to these findings. This study is one of few to successfully leverage bulk CSF RNA-seq data to define biologically plausible mechanisms in neurologic disease.^6,16,17^ Unlike the robust field of host-based bulk transcriptomics in blood^18–20^ and respiratory fluids,^21–23^ the paucicellular nature of CSF severely limits biomass for transcriptomic profiling, posing technical challenges. Our findings confirm that bulk CSF RNA-seq, when paired with rigorous cohort refinement, is an effective method for understanding host response pathways and offers a viable alternative to more complex and costly single-cell sequencing and proteomic approaches.

Owing to the inherent limitations of current TBM diagnostics, most TBM treatment trials recruit fewer than 50% of patients with microbiologically confirmed (i.e. definite) TBM.^9,24–26^ By including patients who do not have TBM, or may have an untreated co-infection, mechanistic/basic science studies are likely to produce misleading results, and clinical trials are inherently biased towards the null hypothesis and therefore less likely to identify novel treatment regimens with therapeutic benefit. Similar to our previous work,^6^ we again demonstrated that mNGS re-classified patients after identifying *Mtb* and/or occult co- or alternate neurologic infections. Here, CSF mNGS reclassified 11% and 12% of participants with probable and possible TBM as definite TBM, respectively, and identified alternate infections in 9% and 12% of these groups. Clinically significant coinfections excluded 5% of patients with definite TBM. These results suggest that other TBM clinical and observational studies (and perhaps more broadly, the Uniform Case Definition for definite TBM)^12^ should consider including CSF mNGS or other broad-based diagnostic testing approaches as a strategy to improve diagnosis and to avoid biasing TBM research results.

### Adaptive immunity in TBM survival

Our transcriptomic analyses revealed a clear pattern in which **most** survivors exhibited robust upregulation of adaptive immune signaling pathways, particularly those involving T-cell and B-cell responses, though as discussed below, important heterogeneity exists within the cohort. The upregulation of TCR signaling components^27–30^ suggests that survivors maintain T-cell signaling activity despite severe HIV-associated immunosuppression. A significant component of the TCR response is likely driven by CD8+ T-cell signaling, as demonstrated by the strong upregulation of class I MHC signaling pathways.^31^ Upregulation of *IL7* (which aids in T-cell homeostasis and survival),^32^ *CD27,* and *IL12RB1* further support the importance of preserved T-cell function in TBM survival.^33–35^ These findings confirm the well-established role of the T-cell response to *Mtb* that has been described across various research models of TBM and clinical data.^36–38^

Unlike T-cell responses, B-cell responses to *Mtb* are poorly understood and historically thought to play a much less significant role.^39,40^ Our transcriptomic analysis, however, demonstrated that B-cell signaling components, including *BCR*, *CD81*, and *PLCG2* were upregulated among TBM survivors. These specific components may reflect upregulation of the antigen presenting role B-cells play in coordination with T-cells, rather than an independent humoral response given the lack of upregulation of genes associated with plasma cell differentiation and immunoglobulin production.^41–43^ However, there likely is also a role for the humoral response, as demonstrated by recent data demonstrating that the CNS-compartmentalized antibody response in TBM can modulate disease severity.^44^ Given that the CSF samples utilized in our analysis were obtained at the time of initial presentation, it is possible that this may have preceded the development of a robust humoral response, and thus longitudinal transcriptomic and antibody-based analyses may be of further utility in understanding the nuances of the anti-*Mtb* humoral response.

### TNF signaling and innate immune balance

Survivors also demonstrated coordinated upregulation of key innate immune signaling pathways, including TNF, a critical signaling molecule of the innate immune response produced by activated macrophages and microglia, along with downstream NF-kB signaling components *CARD10*, *UBE2N*, and *TRAF2*.^45,46^ TNF signaling is known to be an important regulator in *Mtb* control through granuloma formation and other mechanisms, as perhaps best illustrated by the increased risk of TB reactivation in the setting of TNF inhibitors such as infliximab and adalimumab.^47–49^ Furthermore, the *antagonistic* pathway IFN-γ-activated inhibitor of translation (GAIT translation pathway) was downregulated among survivors, resulting in upregulation of these microglial/macrophage functions. Although case reports suggest TNF and IL1 inhibitors can ameliorate dysregulated host responses in TBM immune reconstitution inflammatory syndrome (IRIS) and other paradoxical immune reactions^50–52^, our transcriptomic data emphasize that optimal timing and patient selection for these agents requires caution.

In contrast to the strong proinflammatory signals demonstrated by *TNF*, *IL1*, and T- and B-cell receptor responses, TBM survivors demonstrated downregulation of the neutrophil chemoattractant, *CXCL1*,^53^ the complement pathway genes *C4A* and *C4B*,^54^ and granzyme A signaling.^55^ While neutrophils, complement fixation, and granzyme-mediated cytotoxicity are important for pathogen control, excessive activation could exacerbate cerebral edema and neuronal injury in TBM through generation of reactive oxygen species, increased vascular permeability and excess inflammatory cell recruitment.^54–56^ The downregulation of these specific immune system components in survivors may reflect a protective mechanism that limits neutrophil-mediated and other forms of cytotoxic CNS tissue damage, while preserving more effective and targeted adaptive and innate immune responses. This is supported by whole blood transcriptomic analyses of patients with TBM, which demonstrated that over-activation of neutrophil-mediated inflammation was associated with increased three-month mortality,^19^ and TBM IRIS in HIV-associated disease.^57^ Clinical trials for CXCL1/CXCR2 inhibitors are underway in other infectious and inflammatory diseases,^58^ and complement inhibitors have been approved for immune-mediated diseases including ANCA-associated vasculitides.^59^ Unlike glucocorticoids,^10^ these targeted agents could preserve beneficial adaptive immune responses while selectively downregulating neutrophil-driven innate immune responses. Given the high prevalence of vasculitic stroke in TBM,^60,61^ further investigation of complement inhibitors is particularly warranted.

### Hypoinflammatory phenotype and implications for risk stratification

While the overall pattern among survivors suggests balanced immune activation, the unsupervised hierarchical clustering analysis revealed a distinct hypoinflammatory phenotype characterized by global downregulation of both innate and adaptive immune pathways and significantly elevated mortality. This phenotype underscores that although excessive inflammation can be detrimental in TBM, as evidenced by the downregulation of neutrophil and complement pathways among survivors in the overall cohort, insufficient immune activation may be equally or more harmful. Within this cluster, survivors demonstrated preserved expression of TNF, interferon-gamma, and IL-1 signaling pathways, suggesting a minimal threshold of immune signaling below which survival becomes unlikely even with appropriate antimicrobial therapy. Clinically, this hypoinflammatory phenotype may help explain the lack of definite mortality benefit observed with adjunctive corticosteroids in HIV-associated TBM,^9,10^ as these patients would be expected to experience harm rather than benefit from further immune suppression. The lower CSF lactate in this subgroup suggests that if validated in larger cohorts, routine CSF lactate measurement could help identify these high-risk patients who may require alternative treatment strategies and more intensive monitoring.

### MAPT regulation and neuroinflammation

Beyond the immune phenotypes identified through clustering analysis, upstream regulator analysis revealed additional novel and unexpected findings including the role of MAPT as an upstream regulator in survivors. MAPT is traditionally associated with neurodegenerative diseases, but its predicted activation in survivors, based on downstream targets such as TNF, IL1A, and MAPK1, suggests a potential neuroprotective or neuromodulatory role in acute CNS infections. This finding represents a previously unrecognized mechanism by which the CNS responds to mycobacterial infection. There are no known drugs that upregulate MAPT (on the contrary, antisense oligonucleotides and monoclonal antibodies *downregulating* tau aggregation and MAPT are under investigation for Alzheimer’s disease), though this may be a potential area of interest for future investigation.

## CONCLUSION

This study has several limitations. The cross-sectional design cannot establish causality between differential gene expression and survival outcomes, and longitudinal sampling could clarify whether these expression patterns precede or follow clinical deterioration. CSF-based transcriptomics may not fully capture immune responses occurring in brain parenchyma or meninges, and the paucicellular nature of CSF likely biases results toward robustly expressed genes while potentially missing subtle but important changes. The relatively large BCV values likely reflect this zero-inflated nature of the CSF transcriptional profile. Furthermore, the focus on 14-day survival, while clinically relevant, may not capture longer-term neurologic outcomes. Finally, this study was conducted in two hospitals in Uganda among persons living with HIV, potentially limiting the generalizability of the findings to those without HIV and populations in different geographic settings.

Despite these limitations, we demonstrate that bulk CSF RNA-seq paired with rigorous mNGS-based cohort refinement identified distinct immune mechanisms associated with short-term TBM survival. These findings support investigation of targeted therapies to selectively modulate detrimental innate responses while preserving protective adaptive immunity to improve outcomes in TBM.

## Methods

### Study cohort

Patients presenting to Kiruddu Regional Referral Hospital of Mulago National Referral Hospital in Kampala, Uganda from 2018 – 2023 with known or suspected HIV and concern for CNS infection (such as headache, fever, nuchal rigidity, neurologic deficit, and/or encephalopathy) were screened for study inclusion, as a part of the “Improving Diagnostics and Neurocognitive Outcomes in HIV/AIDS-related Meningitis” study (NCT01802385), a prospective cohort study in Uganda. All patients underwent lumbar puncture during admission with basic CSF studies (cell count, protein, glucose, microscopy, gram stain, cryptococcal antigen). A subset were included in this current study if they met the following criteria: 1) possible, probable, or definite TBM as defined by the Uniform Case Definition for TBM,^12^ 2) anti-TB treatment for 3 or fewer days prior to CSF collection, 3) confirmed positive HIV coinfection, and 4) no known co-infection with any other CNS pathogen. CSF specimens used for these analyses were directly collected into Zymo DNA/RNA Shield collection tubes (Zymo Research; Irvine, CA) and frozen at –80°C within 8 hours of lumbar puncture. The samples were subsequently shipped to UCSF on dry ice for CSF mNGS. This study was approved by Infectious Diseases Institute at Makerere University and the Mulago Hospital Research and Ethics Committee (IRB MHREC1246), University of Minnesota (IRB STUDY00006856), and University of California San Francisco (IRB 13-12236).

Differences in baseline demographics, clinical data, and sequencing parameters were compared between patients who survived to day 14 of hospitalization versus those who died. No participants were lost to follow up. For continuous data, two sample independent t-tests were performed, whereas for binary and categorical data, chi-squared comparisons of proportions were performed.

### CSF metagenomic next-generation sequencing

Total bulk RNA in CSF was sequenced from 133 participants in 6 separate batches. Data from 38 participants was previously published (Sequence Read Archive <u>PRJNA773920</u>).^6^ Total RNA and DNA were extracted from approximately 100μL of CSF for each sample and no-template water controls using the Quick-DNA/RNA Microprep Plus kit (Zymo Research; Irvine, CA). Total RNA was eluted into 22μL of nuclease-free water. RNA-Seq libraries were prepared with the NEBNext Ultra II Directional RNA Kit (E7760; New England Biolabs, Ipswich, MA) according to the manufacturer’s recommendations. Each library was amplified with 18 PCR cycles with incorporation of unique dual-index primers. Quality control was performed on the 4150 TapeStation System (Agilent; Santa Clara, CA), allowing for assessment of average library size and the presence of adapter dimers. Pooling volumes were calculated by performing shallow sequencing on an Illumina iSeq to determine the appropriate equimolar ratios of each sample. Then, the pooled sample libraries were sequenced at the UCSF Center for Advanced Technology on an Illumina NovaSeq 6000 instrument using 150-base pair paired-end sequencing.

mNGS analysis was performed via the open source, cloud-based metagenomics platform CZID.^62^ During the data pre-processing phase, raw fastq files were uploaded and subjected to adapter trimming, and removal of low quality reads, low complexity reads, human reads, and duplicate reads. This is followed by contig assembly and alignment using an indexed version of the NCBI’s GenBank database to identify the source of non-human sequences in the datasets.^62^ The final alignment data were then manually reviewed to determine which reads were correlated with true pathogens. First, the non-template water controls from each run were analyzed to ensure that there were no reads aligning to known pathogens that are *not* commonly seen in CSF mNGS datasets secondary to contamination in the environment or during sample processing (including *Mtb, T. gondii, and C. neoformans*). We then created a background model on CZID using the non-template water controls to establish the relative abundance of reads aligning to organisms in the “contaminome”. Sample alignments were then filtered to only include 1) organisms with known human pathogenic potential, and 2) organisms with a relative abundance greater than the non-template water background model (i.e. Z-score > 1). All potential pathogen reads and contig alignments were manually confirmed via NCBI BLAST.

### CSF differential gene expression analysis

Human transcript counts were generated using a computational pipeline that incorporates quality filtering using the paired-read iterative contig extension (PRICE) computational package (v1.2)^63^ followed by human genome alignment using kallisto (v0.46.1).^64^ In order to account for the pauci-cellularity, low RNA abundance and zero-inflated datasets obtained from CSF, sample weights were calculated prior to differential gene expression analysis. To determine the best fitting model for calculating weight both zero-inflated negative binomial models and standard negative binomial models were estimated using the ZINB-WaVE R package (v1.30.0) after filtering for protein-coding genes only.^65,66^ Combinations of *a priori* suspected confounders or effect modifiers, including demographics (age, sex), CD4 count, sequencing batch, number of non-zero gene counts, and percent guanine/cytosine (GC) content were tested in both zero-inflated and standard negative binomial models. The main comparison of interest, short-term mortality, was not included in the pre-processing model for weights. Akaike Information Criterion (AIC)^67^ values were used to determine not only which model (zero-inflated vs standard negative binomial) better fit the data, but also which combinations of covariates best fit the data. The model with the lowest AIC was a zero-inflated negative binomial model which only included sequencing batch (without other covariates) and was therefore used to generate sample weights for each gene-count.

After applying sample weights, gene counts were filtered to protein-coding genes expressed in >20% of the samples. Differential gene expression analysis was performed using the Deseq2 (v1.48.0)^68^ R package based on 14-day TBM mortality. Differential gene expression (negative binomial regression) models were performed, as well as corresponding biological coefficients of variation (calculated from the square root of the gene-wise dispersion estimates). Differentially expressed genes were defined using a Benjamini-Hochberg adjusted p-value < 0.05 and an absolute log-fold change ≥ 1.5. The data were both manually and computationally filtered (**Supplementary Data 2**) to identify relevant immune genes, and then visualized utilizing the EnhancedVolcano R package (v1.26.0).

To perform the unsupervised hierarchical clustering, the gene count data were filtered to exclude non-protein coding genes and genes with non-zero counts in fewer than 20% of samples. The remaining protein coding genes were then normalized using quantile normalization via the voom function from the limma R package (v3.64.0). The 1,000 protein coding genes with the highest variance, as calculated by applying the variance function across genes, were then utilized to perform hierarchical clustering utilizing Ward’s D2 method on the sample Euclidean distance matrix. The dendrogram was visualized using the ggdendro (v0.2.0) and ggplot2 (v3.5.2) packages, clusters were assigned using the cutree function, and baseline demographic and clinical characteristics were compared across the clusters. Differential gene expression, as described above, was then performed across clusters and within clusters based on mortality.

### Pathway analysis

We performed pathway analysis using the Ingenuity Pathway Analysis (IPA) toolkit (Qiagen; version: 145030503).^15^ All genes that were differentially expressed as defined by a Benjamini-Hochberg adjusted p-value < 0.05 and an absolute log-fold change ≥ 1.5 were included as the input for the analysis. We considered all immunologically relevant pathways that were significant at Benjamini-Hochberg adjusted p-value <0.05 utilizing the built-in pathway filtering methods. In addition, we performed upstream regulator analysis and causal network analysis using the IPA toolkit to identify likely upstream regulators of genes. The activation state of a pathway or gene was calculated based on differentially expressed genes (adjusted p-value < 0.05), and all pathways/causal networks/master upstream regulators with an adjusted p-value <0.05 and absolute Z-score ≥ 2.

## Data and Code Availability

The sequencing data, de-identified clinical metadata, and code will be made available at the time of peer-reviewed publication.

---

**Supplementary Figure 1:**
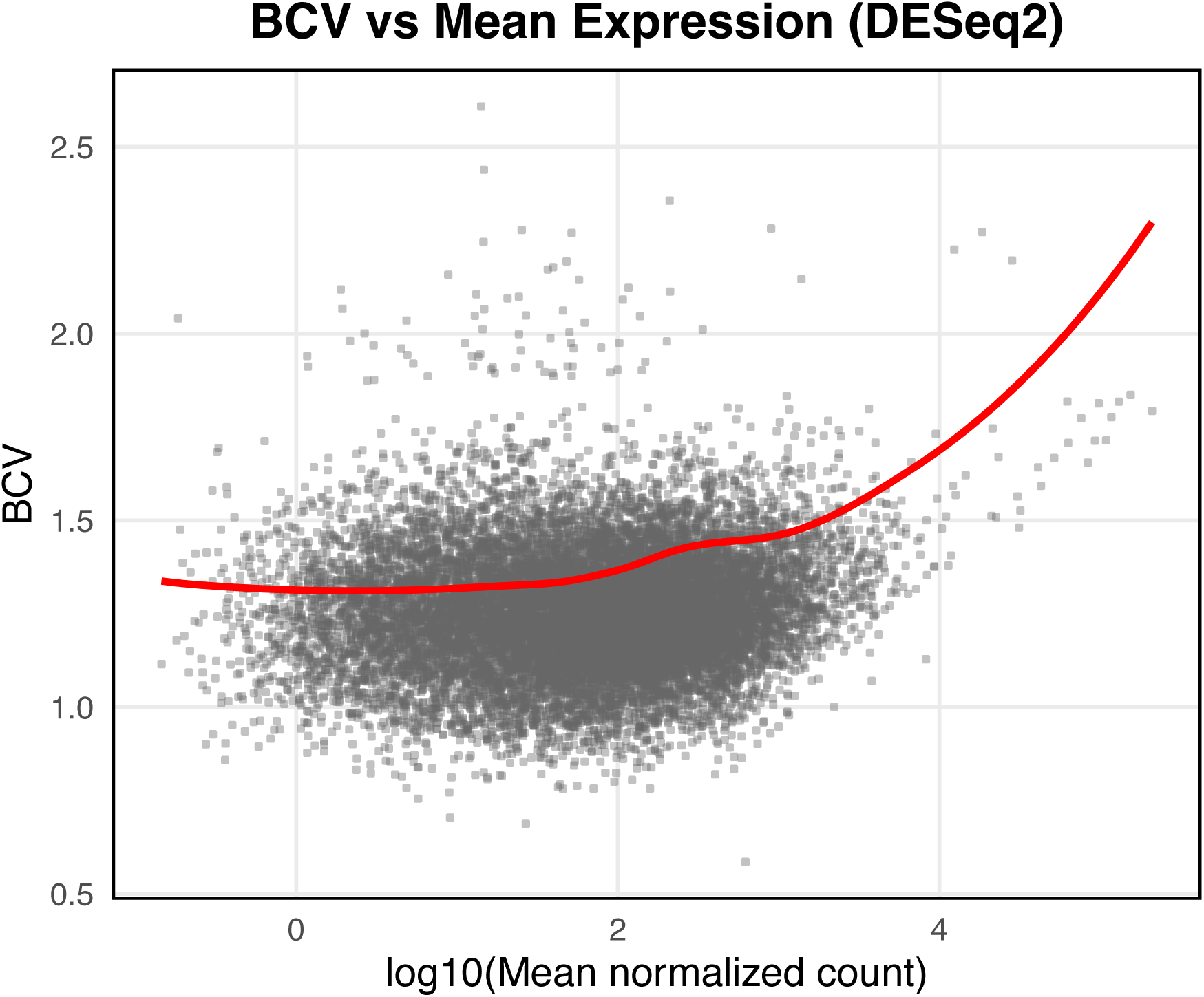
Biological coefficient of variation as a function of mean expression. BCV (square root of dispersion) plotted against log10 mean normalized counts for all genes. The red line represents the fitted dispersion trend estimated by DESeq2.

**Supplementary Table.**
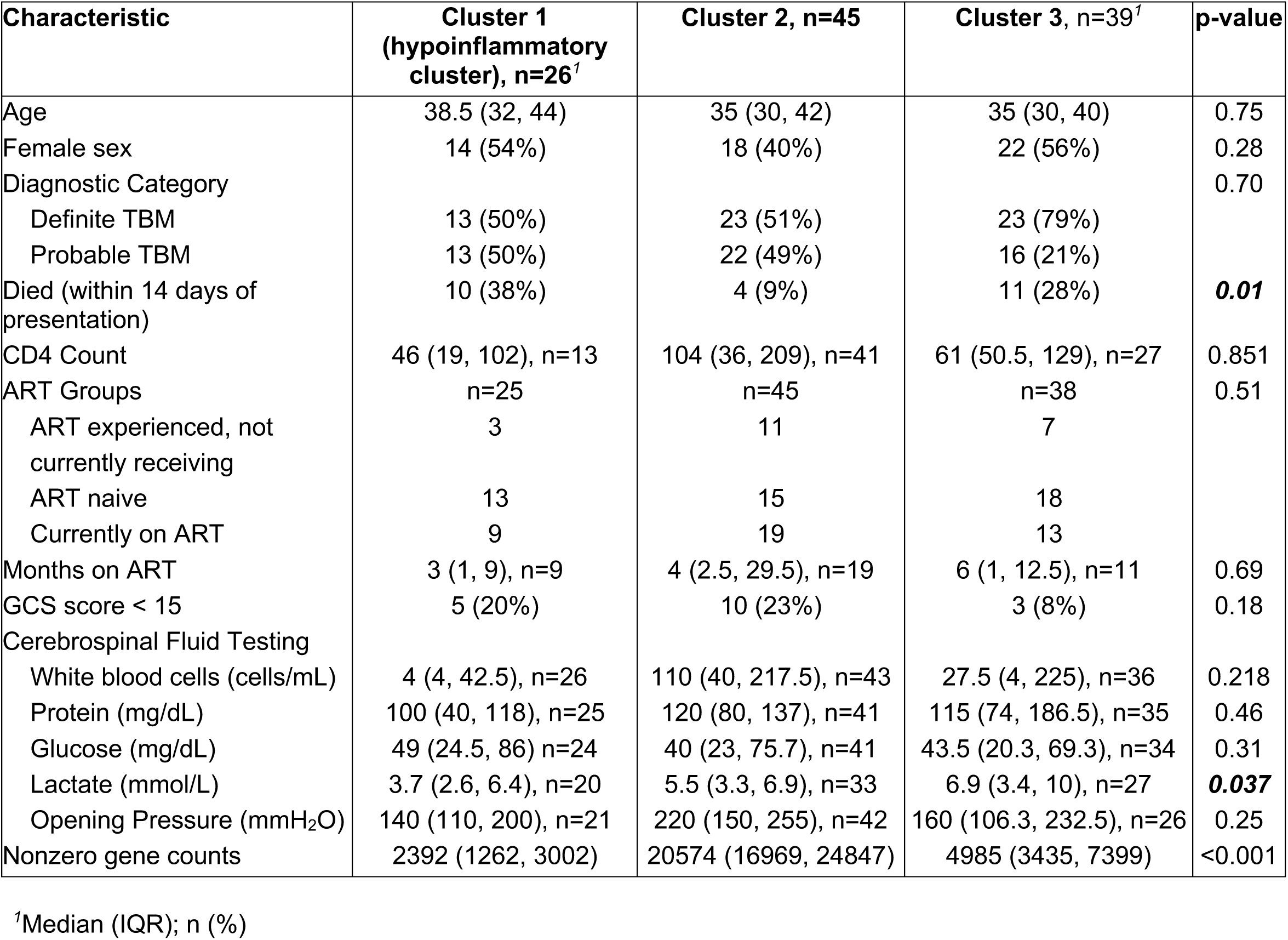

